# Patient-specific finite element modeling of image-derived shell morphoelastic growth: A general framework and application to stented aortic dissection

**DOI:** 10.64898/2026.10.21.26366142

**Authors:** Anna D. Gaffney, Sanjeev Dhara, Charles Davis, Shreya Singh, Žiga Donik, Luka Pocivavsek, Nhung Nguyen

## Abstract

Vascular pathology often manifests in complex shape evolution visualized by longitudinal computed tomography (CT) imaging. Growth and remodeling (G&R) frameworks have been integrated into finite element (FE) modeling to probe the mechanics underlying this progression, extending into patient-specific approaches. Their scalability and translatability, however, are limited by the computational burden associated with heterogeneous, anisotropic growth fields and through-thickness meshing. While existing shell growth formulations can mitigate this burden, they require nontrivial FE implementation especially in commercial softwares (e.g., Abaqus) and their efficacy in patient-specific geometries under complex loading is unclear. In this work, we present a general computational framework to implement shell morphoelastic growth computed directly from registered patient-specific CT images. This shell implementation is validated against solid 3D formulations, allows thickness tunability, and increases compatibility with CT surfaces. We demonstrate the robustness of this formulation through modeling post-operative growth in a patient-specific aortic dissection under stent-based loading. Results first show that complex stent-based loading can deform the shell dissection flap to reproduce post-operative geometry. The outer aortic wall is then given a growth field calculated from registration of 1-month and 5-year post-operative CT images, which is implemented along with stent-based loading and pressurization. Results from this model capture the trends in total and lumen morphological evolution observed at intermediate timepoints. These findings establish a general framework, validated with a patient-specific case study of aortic dissection, with the potential to expand to other complex systems, scale to larger patient populations, and apply G&R to clinical translation.

## 1 Introduction

The natural history of aortic aneurysms and dissections is characterized by progressive tissue degeneration and correspondingly complex changes in aortic size and shape, with often devastating clinical consequences. A substantial body of work has quantified and categorized this shape evolution in patient data through registration of serial computed tomography (CT) images [1–5], while computational modeling offers a complementary route toward mechanistic understanding of the underlying tissue biomechanics [6, 7]. Building on this foundation, growth and remodeling (G&R) frameworks have been used to model aortic tissue biomechanics in-silico and to elucidate G&R mechanisms implicated in disease processes such as hypertension and aneurysmal degeneration [8–11]. These frameworks span a range of mathematical formulations and levels of biological detail, from phenomenological kinematic growth theories [9, 12] to microstructurally resolved constrained mixture models [13–16], trading off computational cost and ease of calibration to patient-specific data. In the interest of broadening applicability and clinical translation, there exists a need for a computational framework for growth that is computationally efficient, remains robust as model complexity increases, and generalizes across patient-specific populations.

Computational growth methods span several formulations, including stress-driven growth [9, 17], damage-driven growth [18–21], and growth fields driven by mechanobiological stimuli such as smooth muscle cell stretch and arterial pulse pressure [22, 23]. Our group has instead pursued a data-driven approach, extracting the growth field directly via registration of sequential CT images of a single patient and computation of local surface area changes [7]. We found that coupling these image-derived, local surface area changes with a finite element (FE) model produces growth predictions that capture patient-specific shape evolution observed in CT images. This suggests that the anisotropic, nonlinear character of the growth field may be sufficient to capture the complex geometric evolution of the anatomy. Despite differences in formulation, each of these approaches confronts the same obstacle: determining and incorporating anisotropic growth fields increase the computational expense of FE simulations, limiting their broader applicability. This burden grows further when such models are applied to CT image-derived, 3D patient-specific geometries rather than idealized cylindrical ones [24, 25]. Adding to the challenge, limited CT resolution introduces uncertainty in evaluating wall thickness [26], which can have a significant impact on both stress calculations and predicted deformation during growth.

Patient-specific modeling has been applied across a range of cardiovascular anatomies, including the aortic valve, abdominal aorta, carotid arteries, and coronaries [27–29]. For FE implementations of growth, these geometries are typically discretized using either 3D solid elements or 2D (shell or membrane) elements, and while several studies have directly compared these formulations in general biomechanics applications [30, 31], the impact on growth modeling is poorly understood. While 3D solid models are widely used for their straightforward implementation and high reported accuracy [32, 33], they are prone to limitations regarding wall thickness and meshing. Tuning wall thickness or specifying spatial variation of thickness, for example, require complete recreation of a 3D model, rendering sensitivity studies and thickness variation costly and impractical. In patient-specific approaches, previous implementations of CT image-derived growth on 3D models also required through-thickness extrapolation of growth fields, further increasing uncertainty [7]. One recent systematic comparison between 3D and shell formulations for non-growth, non-linear vascular biomechanics [31] also found that highly structured through-thickness meshing is required for consistent comparisons in the 3D approach, which is difficult to compute for patient-specific anatomy and leads to computationally expensive small element sizes.

In contrast, shell/membrane models reduce the computational burden and enable effective thickness variation. Shell models are particularly compatible with our data-driven approach to growth modeling, as the CT image-derived growth fields can be directly mapped between surfaces without through-thickness extrapolation. Computation of growth in shell FE models however can require nontrivial implementation, since the out-of-plane stress must be explicitly enforced to zero and the through-thickness strain must be determined [29, 34], which is difficult to satisfy in commercial FE solvers. Additionally, the robustness of such formulations and methodology for implementation in patient-specific geometries under complex physiological loading is lacking. Taken together, the heterogeneity of the growth field compounded by the geometric complexity of resolution-limited, patient-specific CT imaging contribute to the technical and computational demands which limit the scalability and clinical translatability of G&R frameworks.

To illustrate the current limitations of growth modeling, consider the case of aortic dissection, where a tear in the aortic wall leads to the formation of a false lumen for blood to flow between tissue layers. The inner layer pulls away from the outer wall, forming a very thin dissection flap with a complex shape. Modeling the different thicknesses of tissue layers across the entire geometry poses a challenge for 3D modeling, especially when remodeling can lead to a thickening of the flap, even becoming thicker than the outer wall in cases of chronic aortic dissection [35]. The complexity of this case would benefit from a shell formulation for the dissection flap and a separate shell growth formulation of the outer wall to capture the true shape change observed in CT scans. Aortic dissection is typically treated with the deployment of a stent into the true lumen, which applies radial force against the flap and outer wall to redirect flow. This treatment introduces external mechanical stimuli to the aortic tissue, such as pressurization from fluid flow and complex loading from the stent. Without these components, comparison of growth models to post-operative CT data is severely lacking.

In this work, we present a new shell implementation for morphoelastic aortic growth with a heterogeneous, anisotropic growth field extracted from serial CT scans. Its robustness in a patient-specific geometry of an aortic dissection under complex loading from a stent is then evaluated. The structure will proceed as follows: In Section 2, we describe a general computational framework composed of segmentation, image-derived growth formulation, and novel shell FE implementation. In Section 3, we detail the application of this framework to patient-specific aortic dissection under complex loading from a stent through a multi-step process (see Fig. 1). In this application, we first deploy a bare-metal stent into the true lumen of a patient-specific aorta with varying dissection flap stiffnesses. We then calculate an image-derived growth field using CT images from 1 month and 5 years post-operative and model growth in FE with the novel shell morphoelastic implementation. This simulation is conducted in tandem with complex loading and contact conditions from the stent as well as pressurization of the outer aortic wall. In Section 4, we discuss the results obtained from this application, through comparison of total and lumen morphology to CT imaging at intermediate timepoints. Lastly, we discuss the implications of these results and the robustness of our framework in Section 5. To enable widespread adoption and build towards clinical applicability, we also provide example source code and technical implementation guidelines.

**Fig. 1.**
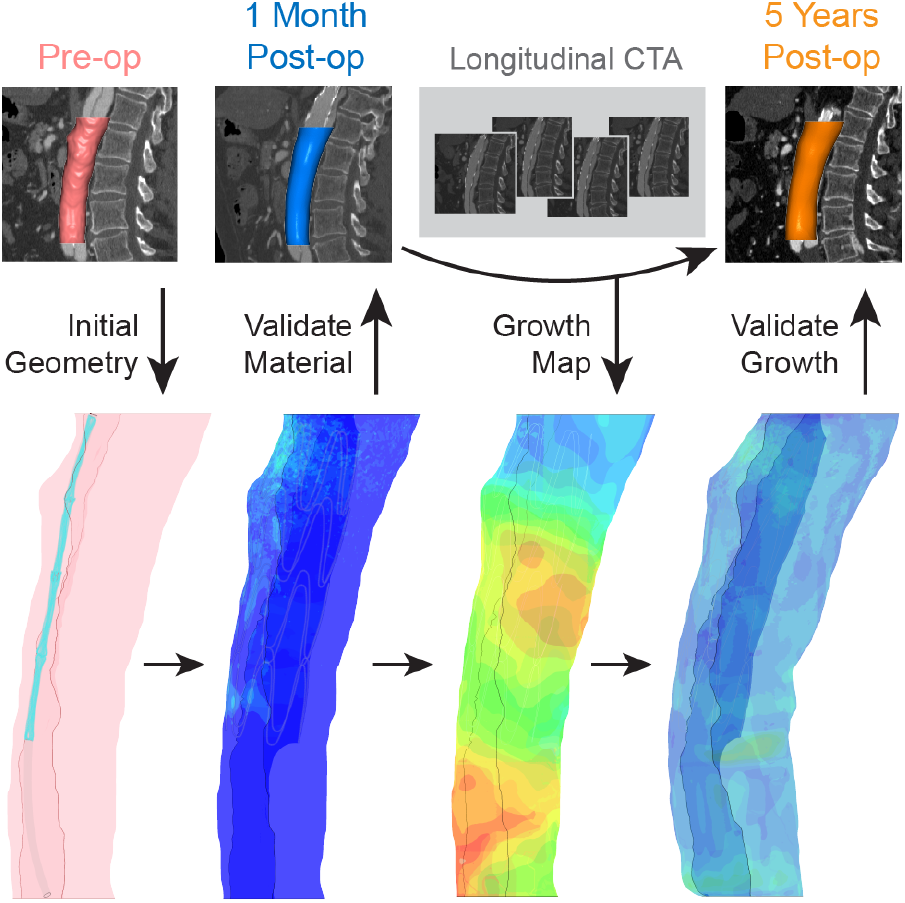
Workflow of application to aortic dissection. From left to right, the CT image-derived pre-operative geometry of the patient’s abdominal aorta (pink) is input into FE simulations as the initial geometry. The dissection flap is also incorporated. Post in-silico deployment of the stent into the true lumen, the resultant deformation is then compared to the 1 month post-op CT image (blue) to confirm effective tissue responses. Then, the CT images from 1 month post-op and 5 years post-op are registered to calculate the heterogeneous growth field, which is mapped onto the deformed in-silico shell geometry and simulated in FE. After in-silico growth modeling, the final deformed geometry is validated via comparison to the 5 year post-op CT image (orange). Additionally, intermediate CT scans (grey) between 1 month and 5 years post-op are compared to intermediate frames in the FE simulation to assess shape evolution.

## 2 General computational framework: Shell implementation of patient-specific CT image-derived heterogeneous growth

### 2.1 Patient-specific geometry

We have previously described a semi-automated pipeline that we use for high-fidelity image segmentation to reconstruct aortas [5, 6]. The segmentation algorithm begins with obtaining CT scans of a patient’s aorta from two timepoints, A and B. Moving forward we will refer to these as aorta A and aorta B. Both scans are segmented using the aforementioned semi-automated algorithm in Simpleware ScanIP, a commercially available segmentation software. This algorithm involves cutting the geometry with axial planes associated with anatomical landmarks and conducting sequential dilation, smoothing, and erosion to minimize measurement artifacts. For the purposes of growth mapping, the smoothed geometry is then exported as a MATLAB surface file for further processing. Additionally, a centerline for the aorta at each timepoint is generated using automated functions within ScanIP and exported as a comma-separated values file (CSV).

### 2.2 Patient-specific image-derived growth field

We have previously developed an image-derived growth mapping algorithm to generate a 3D spatial field of growth rates for application to patient-specific geometries [7]. The first step is to perform a rigid registration between the aorta at timepoint A and B, using the centerline to ensure that the two aortas are in approximately the same physical space. After rigid registration, we then utilize the centerlines to identify corresponding regions of aorta A and B through the unraveling process, described in our prior work [7]. Given each aorta is separated into corresponding regions, we then utilize the coherent point drift algorithm in order to non-rigidly register the datasets on a per region basis. We subsequently utilize the *K*-means algorithm on both aortas to create patches within the regions. Once patching is complete, each triangle on the meshes for aorta A and B is assigned a patch label. For each corresponding patch *k*, we compute the square-root-normalized percent area change:

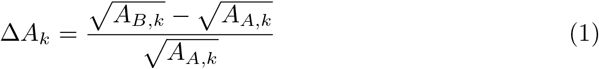

where *A_A,k_* and *A_B,k_* are the total areas of patch *k* in meshes A and B, respectively.

We then compute the growth rate by dividing the area change over a fixed time interval and assign this rate to each triangle *i* belonging to patch *k*:

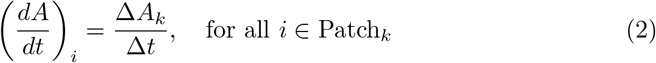

To ensure global continuity of patch growth fields, we apply a *K*-Nearest Neighbors (*K*NN) smoothing algorithm to the patch-wise growth rates. To ensure local continuity across patch boundaries, we also apply a *K*NN smoothing algorithm to the resulting element-wise growth rates, producing a smooth growth field over the surface of mesh A. For compatibility to FE implementation, we export the element-wise growth rates calculated above as a CSV file in the order of the element labels of the aortic wall.

### 2.3 Shell implementation of morphoelastic growth model

Morphoelastic models based on the multiplicative decomposition of the deformation gradient are widely implemented in both commercial FE software and custom codes. In commercial packages, most implementations target 3D solid elements, where integration is comparatively straightforward. Shells are more challenging: plane-stress must be enforced and the through-thickness strain (or growth stretch) must be determined consistently. These requirements are difficult to satisfy especially in commercial FE solvers, where users typically have limited access to the internal stress-update, constraint-enforcement, and linearization routines needed to implement advanced shell G&R behaviors. To date, the majority of studies that utilize shell morphoelastic growth implementations omit critical algorithmic details, such as the through-thickness stretch update to enforce the plane-stress constraint, the evolution of the reference thickness under growth, and the stress update procedure. Moreover, few, if any, studies have addressed the case of a spatially heterogeneous growth field, which is the most relevant formulation to patient-specific, image-derived growth, or compared such results to 3D formulations. These limit reproducibility and broader adoption. Here we present a shell implementation of morphoelastic, image-derived growth for a broad range of applications. This approach not only improves computational efficiency, it also increases the compatibility with the growth field derived from CT imaging in Section 2.2, lessening the uncertainty. FE simulations are performed using the commercial software package Abaqus/Explicit 2025 (Dassault Systèmes Americas Corp., Waltham, MA). Consider the general energy function:

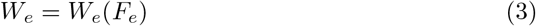

where *F_e_*is the elastic part obtained from the decomposition of the deformation gradient: *F* = *F_e_F_g_*, with *F_g_* as the growth tensor. *W_e_* can be used to describe a variety of material responses, including both general behaviors (e.g., neo-Hookean) and more specific behaviors (e.g., Ogden-Gasser-Holzapfel for fiber-reinforced biological tissue). The growth tensor can be described by multiple growth processes, such as constant growth, stress/strain, or damage-induced growth. We will start with the general implementation and then demonstrate the applications for these individual cases.

The user material subroutine in commercial software such as Vectorized User Material (VUMAT) in Abaqus requires the update of the stress tensors. The second Piola-Kirchhoff stress becomes

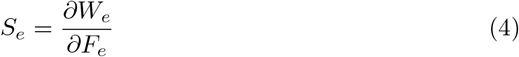

The Cauchy stress needed for implementation is determined through a push-forward process.

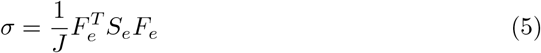

To enforce the plane stress condition in the thickness direction: *σ*_33_ = 0, we use the incompressibility constraint: det(*F_e_*) = 1 to determine the component of the deformation gradient *F*_33_.

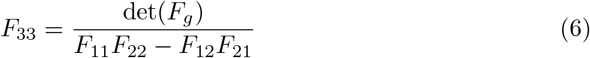

For the shell element, the stress will be modified with the following equations to remove the effect of the hydrostatic pressure:

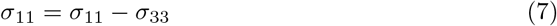

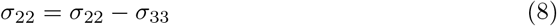

while the through-thickness strain is approximated as follows:

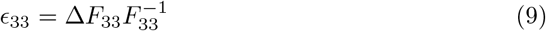

For the morphoelastic growth field implementation in VUMAT that works with both homogeneous and heterogeneous growth fields, we employ the vumatXtrArg subroutine inside VUMAT to access and assign element-wise information. Specifically, to implement the image-derived, heterogeneous growth field, we specify the smoothed growth rates from Section 2.2 into the material definition of the input file and access them using the vumatXtrarg subroutine. The stress and through-thickness strain are then updated, following Equations 7-9. Different forms of the growth tensor *F_g_* can be used. In Appendix A, we present multiple benchmark cases for comparison with the homogeneous *F_g_* = *νI* growth field, which has been the focus of the majority of work in the literature. Specifically, we present the benchmark cases of constant growth (*ν̇* = constant) and stress-induced growth (*ν̇* = *k_ν_*(*ν*)(tr(*M_e_*) *− M_homeo_*). Source codes for these cases are provided for educational purposes and adaptation to future applications. In Appendix B, we further test the difference between shell and 3D implementation using an image-derived, heterogeneous growth field in a patient-specific geometry.

## 3 Application of computational framework

### Post-operative growth of aortic dissection following stent deployment

The robustness of the above framework and implementation approach will be demonstrated in modeling the growth process of an aortic dissection following stent deployment. This clinically important problem is chosen due to its complex loading coupled with a complicated geometry.

### 3.1 Patient geometry segmentation

A patient with a history of chronic aortic dissection had to undergo treatment through stenting of the thoracic and abdominal aorta. The patient received 1 pre-operative and multiple post-operative scans, of which we analyze 1 month and 1, 2, 3, 4, and 5 years post-op. For each of these scans, we utilize ScanIP to trace the aortic wall and dissection flap of the aorta, where we reconstruct a surface model in 3D space. We define the descending/abdominal section by cutting the top boundary at the anatomical landmark of the celiac and cutting the bottom boundary approximately 121 mm descending from the celiac, as seen in Figure 3. Both the aortic wall and dissection flap geometries undergo dilation, smoothing, and erosion processes as described previously [5, 6] and the true lumen (TL) and false lumen (FL) are formed via boolean splitting of the total aortic volume by the dissection flap. For reference, the voxel size for each scan is approximately 0.7x0.7x0.8 mm.

#### 3.1.1 Pre-operative

To incorporate more fine curvature details when applying loads and growth in the FE model, the pre-op geometry is dilated by 4 voxels, mean and median smoothed by 3 voxels, then eroded by 4 voxels to generate the resultant smoothed shape. The flap is then thickened by 1 pixel in the x-y plane to overlap with the aortic wall surface and both geometries are exported as surfaces. These surfaces are imported into the web-based CAD program, OnShape, where the aortic wall is used to clip the flap geometry. This process allows for the formation of 2 open surfaces that can be used for shell implementation in Abaqus: (1) the aortic wall and (2) the face of the dissection flap in contact with the true lumen. Once in the form of open surfaces, the flap is remeshed to ensure all elements, especially those on the clipped edges near the aortic wall, have a near 1 aspect ratio. These surfaces are then exported as STL files and imported into Abaqus for FE modeling (detailed in Section 3.3).

#### 3.1.2 Post-operative

For all other CT scans, a more coarse smoothing is applied to the geometries. These scans are dilated by 4 voxels, mean and median smoothed by 5 voxels, recursive gaussian smoothed with a gaussian sigma of 7 voxels, then eroded by 4 voxels. The dissection flap geometries are dilated by 1 voxel, recursive gaussian smoothed with a gaussian sigma of 2 voxels, then eroded by 1 voxel. For growth mapping, the 1 month and 5 year post-op scans are further exported as surface files. For these geometries we also generate centerlines for the aortic wall geometry utilizing the automated functions within ScanIP, exported as CSV files.

### 3.2 Growth mapping

For implementation of growth after stent deployment in the specified patient, we calculate a heterogeneous growth field via the local area changes of 200 patches from 1 month post-op to 5 years post-op (see Fig. 3). The area changes are divided by Δ*t* = 60 months to calculate the growth rates. For application of this growth map to the pre-op geometry with the stent deployed in-silico, we conducted one additional rigid registration between the pre-op aortic wall mesh used in Abaqus and the 1 month post-op CT image-derived geometry. Using nearest neighbor approximation, each pre-op mesh element is assigned a patch label on the 1 month post-op geometry. The growth field is smoothed over 20 nearest patches, then smoothed over 500 nearest elements for the final growth field shown in Fig 2.

**Fig. 2.**
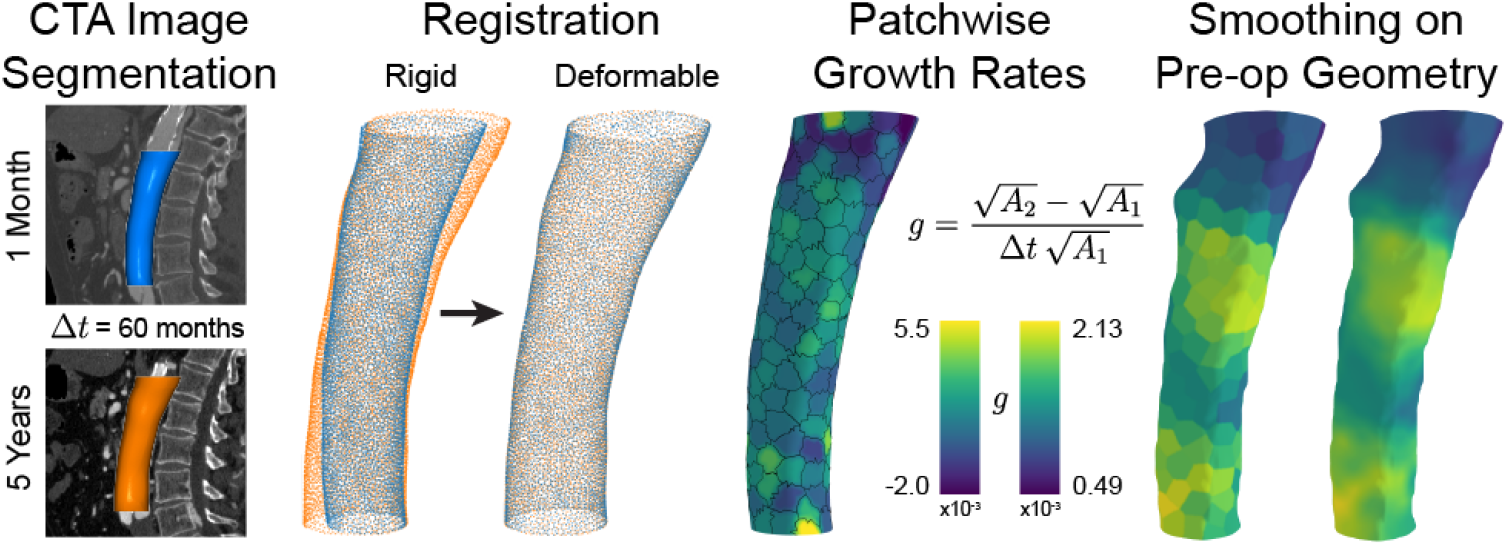
Workflow for calculation of patient-specific, image-derived, heterogeneous growth field. From left to right, CT images are segmented into surface geometries in 3D space at two timepoints. These geometries are registered rigidly at first, then the initial geometry (blue) is deformed to match the final geometry (orange). Sections of these geometries are assigned corresponding patches, and the area change of each patch is calculated into a growth rate (*g*), shown as a heat map. These growth fields are then smoothed globally (patch-wise), followed by locally (element-wise), and finally implemented in the FE simulation.

**Fig. 3.**
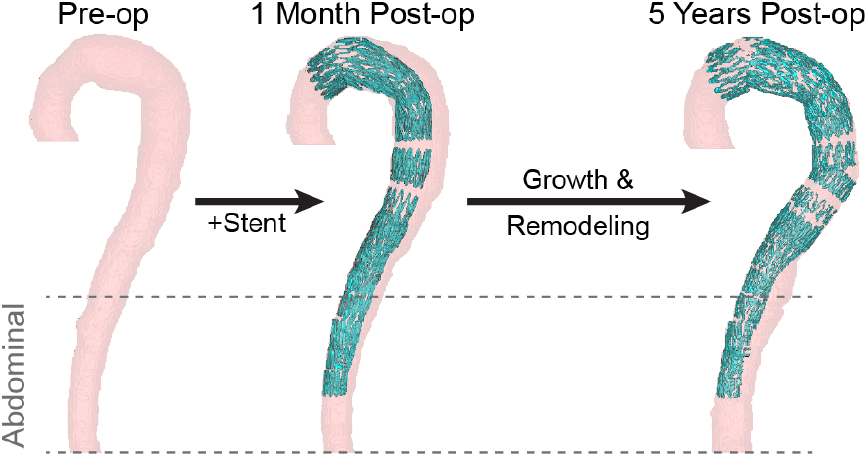
Longitudinal patient-specific case study: aortic dissection. Surface segmentations in 3D space of the CT image-derived aortic geometry (pink) and deployed stent (blue) at pre-op, 1 month post-op, and 5 years post-op. In this work, we model the abdominal portion, as indicated with the grey dotted lines.

### 3.3 Finite element simulations

#### 3.3.1 Assembly and materials

The model assembly is made of 4 parts: (1) aortic wall, (2) dissection flap, (3) stent, and (4) crimper. (1) The aortic wall is derived from the pre-op patient geometry as described in Section 3.1 and is composed of 164933 two-dimensional shell elements (C3, 3-node triangular general-purpose, finite membrane strains) with a thickness of 2 mm and 5 thickness integration points. The shell morphoelastic growth model is implemented through a VUMAT as described in Section 2.3, where the growth rates are calculated as described in Section 3.2, but are not applied until the growth step. Before the growth step, the base material acts as a Neo-Hookean hyperelastic. Before and during growth, the aortic wall is modeled with a *C*_10_ = 0.174 MPa, *D*_1_ = 0.28736

MPa*^−^*^1^, density = 1000 kg*·*m*^−^*^3^. (2) The dissection flap is also derived from the pre-op patient geometry as described in Section 3.1 and is composed of 47541 two-dimensional shell elements (C3, 3-node triangular general-purpose, finite membrane strains) with a thickness of 2 mm and 5 thickness integration points. Note that, while we will mainly focus on the stiffness of the dissection flap as a measurement for chronicity in the following section, due to the shell implementation, we can conveniently study the thickness sensitivity of the flap as well. To mimic the stiffening of the flap, the isotropic hyperelastic material model proposed by Vorp and Raghavan that has been used primarily for aortic tissue in the literature [36, 37] is used. Specifically, we utilize a built-in 2^nd^ order reduced polynomial hyperelastic with a *D*_2_ = 2 *×* 10^6^ MPa*^−^*^1^ to approximate the lack of second-order volumetric response. While the density is kept the same at 1000 kg*·*m*^−^*^3^, the other material parameters are varied to alter flap:aorta stiffness ratios as detailed in Table 1.

**Table 1.** Material properties associated with each flap:aorta stiffness ratio used in this work.

| flap:aorta ratio | $C_{10}$/MPa | $C_{20}$/MPa | $D_1$/MPa<sup>-1</sup> |
| --- | --- | --- | --- |
| 1 | 0.174 | 1.881 | 0.0004 |
| 0.1 | 0.0174 | 0.1881 | 0.004 |
| 0.01 | 0.00174 | 0.01881 | 0.04 |
| 0.001 | 0.000174 | 0.001881 | 0.4 |

In addition to the biological materials, we utilize two parts for stent deployment. (3) The stent is composed of an assembly of four identical single stent rings. The stent ring is designed in OnShape as a sinusoidal curve/wire part, parametrized with 6 equidistant peaks, a ring length of 20 mm, a ring diameter of 36 mm, and a wire diameter of 0.5 mm, approximating an example geometry of a bare-metal stent ring. This geometry is exported as a STEP file and imported as a part into Abaqus. In Abaqus, each ring part is composed of 36 one-dimensional explicit beam elements (B31, 2-node linear beam) with a circular profile of diameter = 0.5 mm. The stent is modeled as a linear elastic approximating nitinol, where Young’s Modulus = 83 GPa, Poisson’s ratio = 0.33, and density = 6450 kg*·*m*^−^*^3^ [38]. Four identical rings are stacked axially on top of each other, with the bottom-most ring rotated 30*^◦^* to be out of phase with the rest. All peak nodes lay on 1 of 5 planes along the stent length. On each plane, all peak nodes are connected via a continuous wire connector segment, acting as the suture thread that holds the bare-metal stent together. These connector segments are given two axial linear elastic behaviors: stiffness = 1 *×* 10*^−^*^7^ MPa under compression (slack) and stiffness = 1 *×* 10^3^ MPa under tension (taut). To model the direct suture connection of the bottom two ring peaks that are in contact, kinematic coupling of translational displacement between connected peak nodes is implemented. For natural deformation of the stent, instead of applying loads or displacements directly to the stent nodes/elements, all loads are applied indirectly through programmed displacement of a crimper, as developed previously [38]. (4) The crimper is designed in Abaqus as a cylindrical tube of length = 125 mm and diameter = 36.5 mm, creating a 0.25 mm gap between the crimper wall and center of the stent beams as allowance for beam thickness. The crimper is composed of 575 two-dimensional surface elements (SFM3D4R, 4-node quadrilateral, reduced integration) with a density of 7850 kg*·*m*^−^*^3^. Surface elements are like membrane elements, defined as surfaces in space with no inherent thickness or stiffness and act as a rigid body. Structured homogeneous meshing allows for consistent loading along the stent.

#### 3.3.2 Boundary conditions and interactions

In this work, two FE simulations are detailed. The first simulation models the in-silico deployment of the bare-metal stent into the patient-specific aortic geometry. This simulation has three steps: (1) crimp the stent, (2) bend the stent into the aortic true lumen, and (3) expand the stent into contact with the aortic wall and dissection flap. To optimize computational load, the final step is conducted in a separate simulation, using the deformed state from the end of step 3 of the previous simulation. In this second simulation, the final step (4) implements the shell morphoelastic growth described in Section 2.3. Figure 4 visualizes this pipeline.

**Fig. 4.**
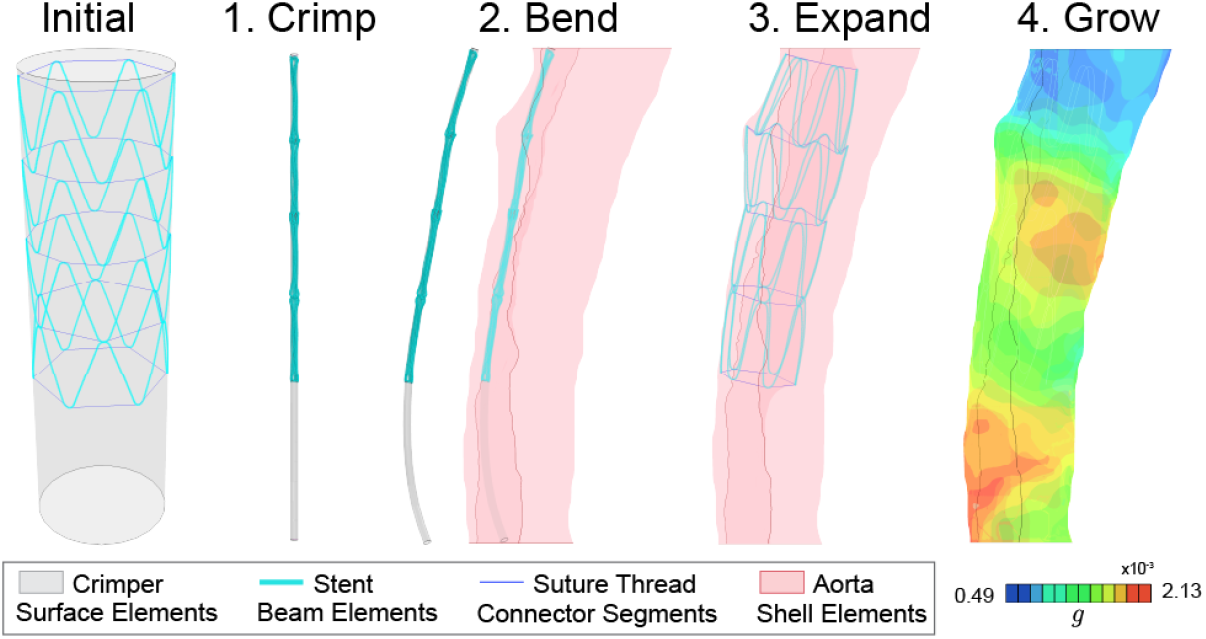
Workflow for finite element simulations. The initial geometry of the crimper, stent, and suture thread. (1) In the crimp step, the crimper radially deforms the stent. (2) In the bend step, the crimper bends the stent to follow the centerline of the true lumen. (3) In the expand step, the crimper allows the stent to expand inside the true lumen until it comes in contact with the aortic wall and dissection flap. (4) In the grow step, the stent remains in contact, applying forces, while the image-derived, heterogeneous growth field is applied to the aortic wall.

In the crimp step (1), the goal is to homogeneously decrease the diameter of the crimper until it is below the minimum diameter of the aortic true lumen geometry, which was measured to be approximately 6 mm. With a starting diameter of 36.5 mm, each node in the crimper is given a displacement boundary condition of 17.25 mm radially inward, to reach a crimped diameter of 2 mm. Cartesian coordinates are used to allow ease of displacement calculations in later steps. General contact interactions are only applied to the stent and the crimper, with penalty tangential behavior (friction coefficient of 0.5) and hard contact normal behavior. This allows the crimper to act as a tool to collapse the stent into a diameter of 2 mm.

In the bend step (2), the goal is to bend the crimper to align with the centerline of the aortic true lumen geometry and we adapt our previously published workflow here [38]. The crimper is meshed into 26 rings of nodes. As such, the centerline for the aortic true lumen as derived from ScanIP is split into 26 points, with cartesian positional coordinates and tangent vectors. To calculate the displacement vector required to align the crimper with the centerline, each ring of crimper nodes is considered a planar geometry. That plane is then subject to displacement and rotation to match the position and direction of the associated centerline point. The displacement vector for each node is then calculated to maintain original planar geometry.

In the expand step (3), the goal is to expand the crimper to allow controlled, yet natural expansion of the stent into the aortic true lumen geometry. Utilizing the same planar geometries from the previous step, the radial displacement of each crimper node outward 17.25 mm is calculated through subtraction of the centerline coordinate from the crimper node coordinate. The previously defined general contact interactions are continued between the stent and the crimper, with added interactions between the stent and the inner aortic surfaces. This allows the crimper to slowly expand, passing through the aortic shell elements, until the stent is fully in contact with the aortic surfaces and no longer in contact with the crimper. Additionally, the top and bottom boundaries of the outer aortic wall are fixed in all directions, while the top and bottom boundaries of the dissection flap are fixed only in the z (axial) direction.

In the grow step (4), the deformed state of the stent and aorta from the end of the previous step are imported as the initial state of a new simulation. The previously defined general contact is continued between the stent and the aortic shell, as well as the previously defined boundary conditions for the edges of the aorta. While growth is applied via material property, as detailed in Section 2.3, an additional pressure load of 0.01 MPa is applied normal to the inner surface of the outer aortic wall to account for pressurization associated with blood flow and fortify structural integrity of the internal cavities.

### 3.4 Post-processing

#### 3.4.1 In-silico deformed geometry

In-silico deformed geometries are exported from Abaqus as separate aortic wall and flap STL files and imported into ScanIP for processing and analysis. The aortic wall is closed and converted into a mask. The flap surface, which is the side that is in contact with the true lumen, is then translated towards the false lumen 1 mm (positive y direction), and converted into a mask using a robust algorithm, which thickens the flap to a thickness of approximately 2mm, allowing for the original flap surface to align with the true lumen side of the thickened flap mask. The true lumen and false lumen are thus formed via boolean splitting of the total aortic volume by the dissection flap.

#### 3.4.2 Measurement of lumen and total volume

To compare between deformed computational results and CT geometries, measurements of volumes and areas are conducted in ScanIP. Deformed computational geometries are imported and processed as previously described. The following measurements utilize mask statistics in ScanIP: Total volumes for the entire cavity (true lumen, false lumen, and flap) are measured, as well as each lumen separately. For lumen area ratios, surface areas of each lumen mask at designated axial slices are measured.

## 4 Results and discussion

With the longitudinal CT imaging spanning from pre-op to 5 years post-op, the dynamic evolution of aortic shape, size, and lumen ratio can be assessed directly. To the best of our knowledge, there is currently no in-silico patient-specific G&R model that captures and is validated against CT image-derived lumen area changes over time. In the following section, we show that utilizing our established growth mapping techniques, we can extract and incorporate the heterogeneous growth field to simulate post-operative remodeling of the outer wall. The developed shell implementation of morphoelastic growth exhibits great strengths outside of the expected benefits of shell implementation (e.g., simple meshing requirement and lower computational costs). These strengths include (1) its ability to perform robustly under complex loading from stent deployment against both the dissection flap and aortic wall, (2) simplification of the growth mapping process, since there is better compatibility between surface-based CT scans and shell elements, and a through-thickness growth field does not need to be inferred or prescribed to the model, and (3) while here we focus on sensitivity to flap:aortic wall stiffness ratios, the procedure is also effective in studying sensitivity to thickness. Our results capture the trend of true lumen:false lumen (TL:FL) evolution in CT scans at intermediate timepoints, even though only the 1 month and 5 year post-op scans were used in calculating the growth field. This demonstrates the potential utility of this approach in understanding the aortic growth captured in routine imaging.

### 4.1 Stent deployment induced shape change

To compare in-silico growth to patient CT imaging, the deformation of the flap due to the radial forces of the stent must align with CT data. For this purpose, the stent is deployed in-silico while varying the stiffness of the flap compared to the aortic wall as follows: 1, 0.1, 0.01, 0.001. Stiffness of the aortic dissection flap has been linked previously to dissection chronicity [35], with increasing stiffness as a result of post-dissection remodeling or fibrosis. It is first found that, in agreement with past literature [38], decreasing flap stiffness leads to a larger flap deformation and an increase in the size of the true lumen, as seen in Figure 5a. We can then examine the impact of flap stiffness on the stress distribution on the aortic tissue. Figure 5a shows the Von Mises stress distribution after deployment. For all models, the stress is concentrated where the stent beams make contact with the aortic wall. Higher stress levels are associated with higher flap stiffness due to the increased constraint on the stent’s deformation, applying more radial force on the aortic wall.

**Fig. 5.**
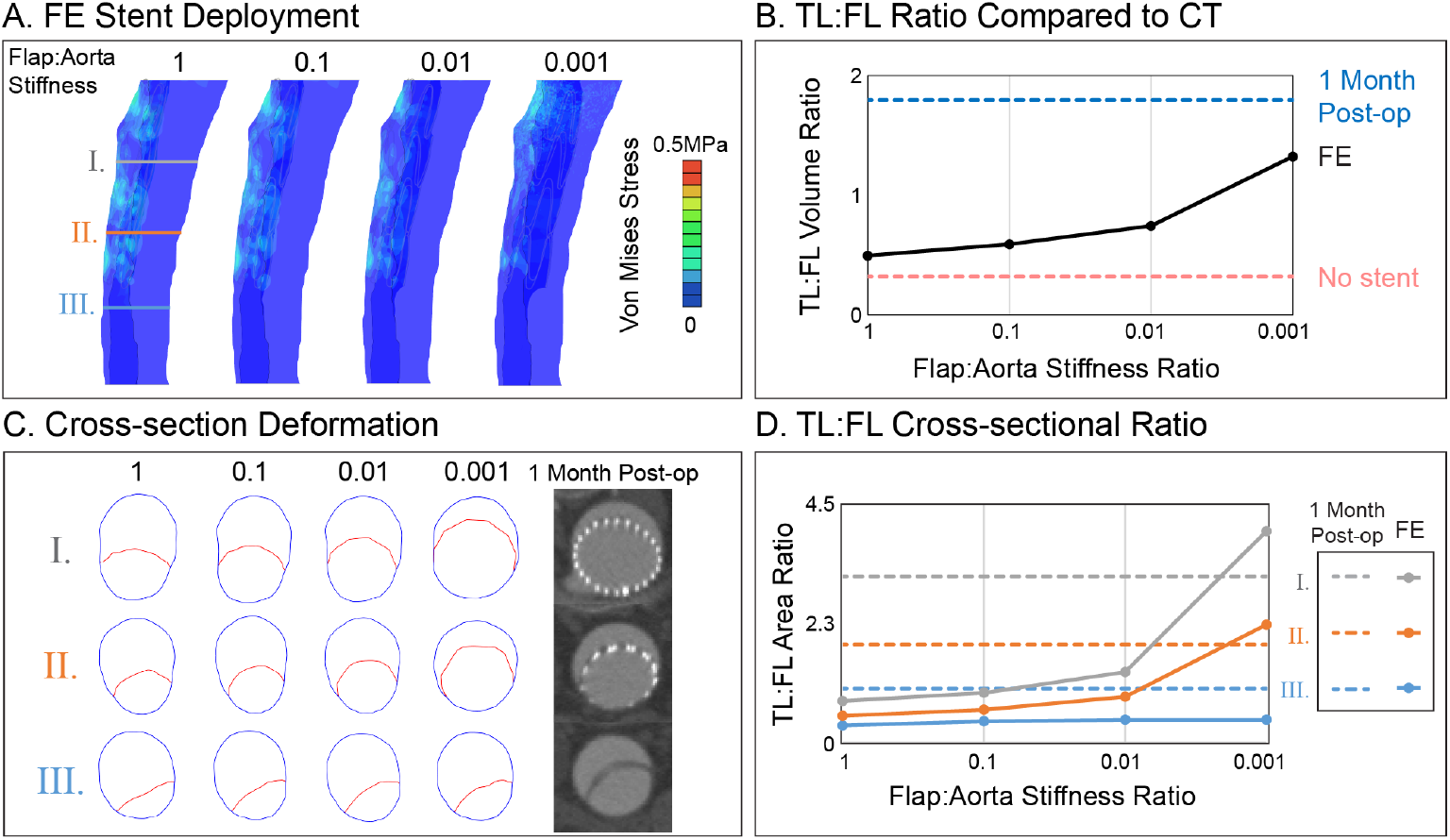
Shape change due to in-silico stent deployment. (A) Deformed geometries after stent deployment for FE models with varying flap:aorta stiffness ratios. The Von Mises stress profile from the stent loading is shown. On the leftmost aorta, three axial slices are defined (I, II, III). (B) TL:FL volume ratio of deformed FE models plotted for varying flap:aorta stiffness ratios. Reference TL:FL ratios for the initial pre-op and the 1 month post-op CT geometries are shown as dotted lines. (C) Axial cross-sections for the outer aortic wall (blue) and dissection flap (red) of deformed FE models at points along aorta as shown in A. The associated cross-sections for the 1 month post-op CT scan are shown to the right. (D) TL:FL area ratio of deformed FE models plotted for varying flap:aorta stiffness ratios, at the 3 points along the aorta as shown in A and C. Reference TL:FL ratios for the 1 month post-op CT scans as shown in C are depicted as dotted lines.

To quantitatively determine the impact of stent deployment on flap deformation and compare to clinical data, the ratio of TL to FL volume is measured before and after the stent is deployed in-silico. Before the stent is deployed, all models begin with a TL:FL volume ratio of 0.3:1. From the CT data at 1 month after stent deployment, the TL:FL volume ratio increased to 1.8:1. For in-silico stent deployment, with decreasing flap stiffness, the TL:FL volume increases (Fig. 5b). In this patient’s case, the flap:aorta stiffness ratio of 0.001 is shown to have the closest agreement with the 1 month post-op CT, with a TL:FL volume ratio of 1.3:1. While this value approaches the clinical value, the following exploration of how the TL:FL ratio changes axially indicates why the flap:aorta stiffness ratio of 0.001 is the best choice for an accurate material model. To explore the impact of the stent loading on flap displacement along the aorta, we extract cross-sectional deformed geometry for regions towards the beginning of the stent (I), end of the stent (II), and where there is no stent (III) (see Fig. 5a). From this view, the similarity in flap deformation between the 0.001 stiffness ratio and 1 month post-op CT geometry becomes clear, specifically in the regions with the stent (I, II). In the region without the stent (III), this view shows how little impact the stent loading has on regions further down the aortic geometry in-silico. In contrast, the 1 month post-op CT image shows a much more even TL:FL ratio even where no stent is present. We hypothesize this is due to the additional pressurization in the TL due to the increased cross-sectional area from the stent which allows for more blood flow. These findings are quantitatively corroborated in Figure 5d, where each of the three cross-sectional regions for the four in-silico cases are compared to the 1 month post-op CT. For regions with the stent, the 0.001 stiffness ratio model has a slightly higher TL:FL ratio, while the opposite is true in regions without the stent.

The proposed shell formulation allows for significant deformation of the dissection flap, which is required to capture the 1 month post-op CT geometry for this patient. While only the flap:aorta stiffness ratio is tested in this section, this shell implementation enables variation in flap thickness and outer wall thickness separately, or even variation of the thickness spatially across the surface. Tuning these thicknesses will impact the stress distribution and deformation. Therefore, future work may utilize flap thickness as another method to alter the material response and capture patient-specific deformations.

### 4.2 Growth with stent loading

In the previous section, we determine that a 0.001 flap:aorta stiffness ratio leads to the geometry that most closely aligns with the 1 month post-op CT data. From this model, we utilize the deformed geometry, stress profile, and material properties as the base system to simulate post-operative growth. As described in Section 3, we conduct growth mapping from the 1 month post-op CT geometry to the 5 year post-op CT geometry. After smoothing iterations, we generate a spatial growth map (Fig 2) that is applied to the deformed FE model from the previous section (Fig 4). For this patient specifically, their growth is distributed highly heterogeneously, with 2 regions of peak growth and one region of minimal growth. The growth rates calculated here are for 60 months (or 5 years), consider the start of the simulation as time of operation, and are applied constantly for temporally linear growth. To avoid unrealistic deformations and approximate fluid flow, a small pressure load is applied to the aortic wall during this step.

We first look at the overall size and shape change as a result of the implemented growth field. In Figure 6a, the size of both the FE model and patient geometry gradually increases over time. This is quantified in Figure 6b, with the total volume of the aortic wall geometry increasing at a very similar rate. This result is nontrivial and indicates that the growth of this patient’s aortic wall can be captured with a temporally linear growth rate and the designated spatial growth map. In addition to the size of the aortic wall geometry, we also take note of the shape of this geometry. In the second row of Figure 6a, it is seen that the axial curvature of both the FE model and patient geometry gradually increases over time. In the FE simulation, this even leads to a localization of high Von Mises stress in the most curved/bent region of the deformed geometry.

**Fig. 6.**
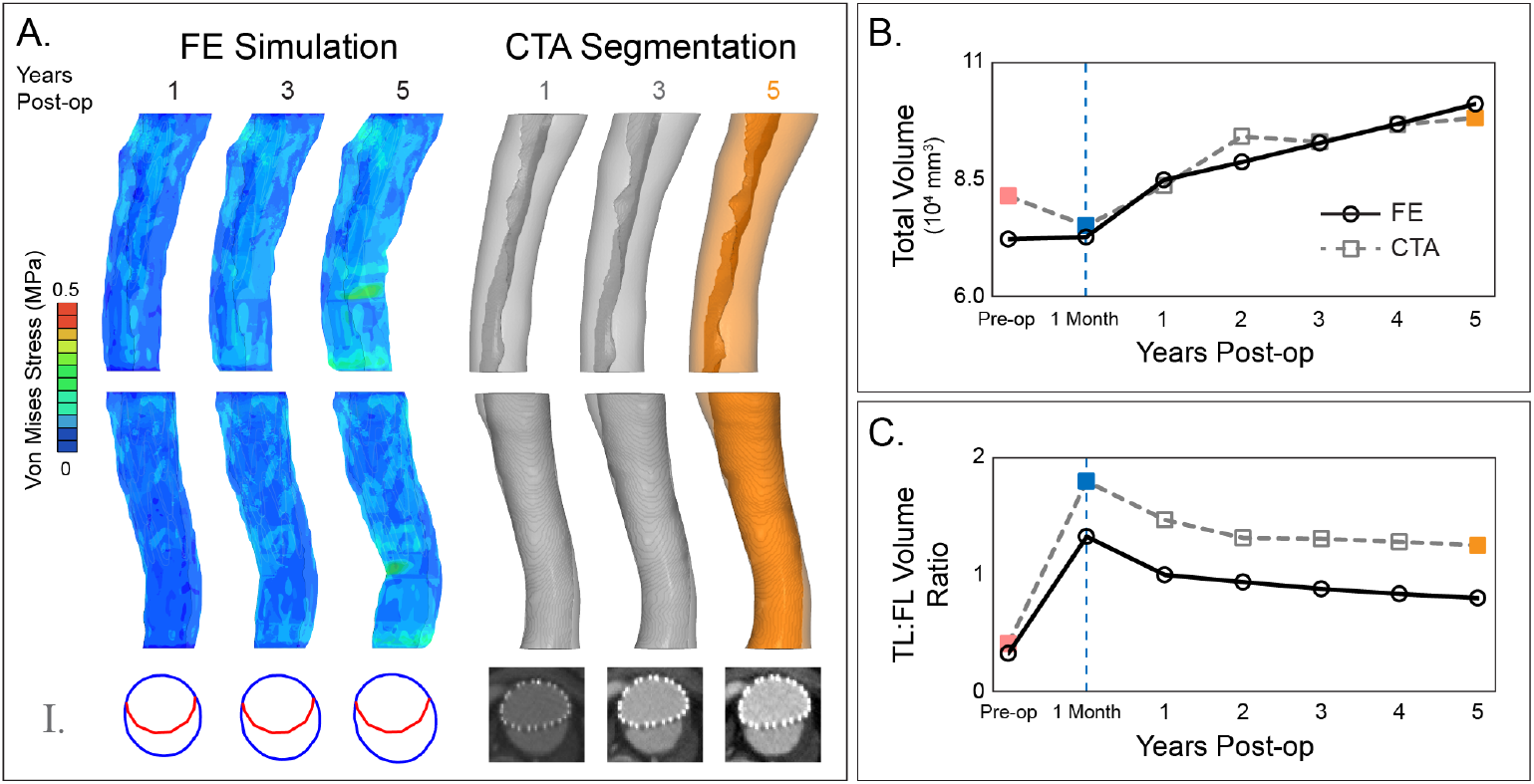
Shell implementation of image-derived growth with complex loading. (A, left) Deformed geometries throughout the growth process in FE. The Von Mises stress profile from the growth and pressurization is shown. Below are corresponding axial cross-sections of the aortic wall (blue) and dissection flap (red) for the slice I from Fig 5a. (A, right) Segmentations of post-op CT scans at three timepoints. The intermediate geometries (1 & 3 years post-op) are not included in the simulation workflow. The 5 year post-op geometry (orange) was utilized to calculate the growth field. Below are corresponding axial cross-section CT scans for slice I. (B) Total volume of the space enclosed by the outer aortic wall for all timepoints throughout the growth process. (C) TL:FL volume ratios for all timepoints throughout the growth process. *Note: FE = solid, black, circle; CT = dash, grey, square*.

While this growth mapping is shown to capture the post-operative growth of the outer aortic wall, we are most interested in capturing the change in TL:FL ratio during this growth. As seen in the first row of Figure 6a, the true and false lumens of the patient’s aorta increases over time. In Figure 6c, we quantitatively measure the TL:FL volume ratio for the patient CT scans (dotted) and FE simulation (solid) as a function of years post-op. Interestingly, the trend in the patient’s TL:FL volume ratio evolution is also captured with our model that incorporates growth mapping and consistent complex loading from the deployed stent. We attribute the systematic gap between the CT and FE TL:FL volume ratio to the region of the aortic geometry without a stent, which continues to exhibit a higher TL:FL ratio in the CT in comparison to the FE model.

While the evolution of the TL:FL ratio is in good agreement, we note that the shapes of the two lumens are not in complete agreement. As seen in the third row of Figure 6a, one representative cross-sectional deformed geometry showcases how the flap remodels and forms a flatter shape in the years post-op. Our shell implementation can be pushed further in the future to address this discrepancy, such as varying thickness over time or incorporating spatial changes of thickness to impact the geometry and shape the dissection flap to align more with the CT data.

## 5 Conclusions

As noted in the Introduction, despite the tremendous efforts in modeling growth and remodeling processes, the associated technical and computational challenges require new computational development to address complex conditions and analyze large numbers of patients. In the case of aortic dissection, modeling growth in a patient-specific geometry can introduce complexity through the varying thickness of the dissection flap, uncertainty in the geometry, and heterogeneous growth fields that may be extracted from CT image-derived surfaces. These components can further increase the computational cost and make analysis of large datasets impractical. In this work, we present a generalized computational framework to segment patient-specific geometries, calculate CT image-derived growth fields, and implement morphoelastic growth in a shell formulation. The proposed novel shell formulation lowers the computational cost of growth modeling and allows for wall thickness tunability, which is necessary to address uncertainty due to the CT resolution limit. It also enables more effective mapping of patient-specific growth fields from CT surfaces to shell surfaces in FE. We validate the accuracy of this implementation against growth in existing solid 3D formulations and shell formulations (Appendix A), noting the highest accuracy in geometries with low ratio of wall thickness to radius. Different formulations of growth are also presented, such as homogeneous and stress-driven growth, showcasing its adaptability to other applications.

To further demonstrate the effectiveness of the developed method, we then apply the proposed methodology to growth in a patient-specific geometry with aortic dissection, including additional complexity through the dissection flap, bare-metal stent loading, and pressurization as a proxy for fluid flow. We first create a CT image-derived shell model from the pre-operative patient geometry, including the outer aortic wall and dissection flap. We determine the effective stiffness ratio between the dissection flap and aortic wall through in-silico stent deployment and comparison of lumen volumes to post-operative CT imaging. Our results indicate this patient exhibits a dissection flap that is approximately 0.1% the stiffness of the aortic wall (Fig. 5).

We then calculate an outer aortic wall growth field through registration of 1 month and 5 year post-operative patient geometries, followed by calculation of local area changes. We implement this growth field utilizing the novel shell growth formulation in tandem with stent loading and pressurization. We compare the resultant size and shape evolution to CT image-derived lumen volumes at intermediate timepoints between 1 month and 5 years and find good agreement (Fig. 6). These results suggest this model captures the longitudinal evolution of size and shape for an aorta under complex loading, simply through modeling image-derived growth of the outer aortic wall. The computational stability during this complex application showcases the robustness of our proposed shell growth implementation and feasibility to utilize this framework for other complex or computationally expensive systems.

The main limitation of this shell implementation of growth is the requirement of low ratio of wall thickness to radius for high accuracy simulations, as shown in Appendix A. While the thickness to radius ratios for vasculature are typically quite low, this limitation should be noted. In our application to a patient-specific aortic dissection, this case study is limited to one patient. With the backing of these validated results and the developed methodology, this workflow can now be utilized to study larger sets of patients with longitudinal CT imaging. In this work, for simplicity, we only test one thickness for both the aortic wall and the dissection flap. Spatially varying the thickness throughout or between either of these surfaces could enrich this work further, and can be incorporated with this shell implementation without any reconstruction. Lastly, this work does not utilize a multilayer representation of the aortic wall, the addition of any pre-stress or residual stresses, or remodeling of the dissection flap over time. Future adaptations of this shell implementation may incorporate these components into the growth formulation to connect to mechanobiological processes.

Overall, this work fills an existing need for a computational framework for growth that is computationally efficient, is robust under increasing degrees of complexity, and generalizes across patient-specific populations. Our proposed shell morphoelastic growth FE implementation is validated against solid 3D formulations, allows thickness tunability, effective mapping from CT to FE, and allows for exploration of growth under complex loading. Our FE analysis of patient-specific aortic dissection shows this robustness and offers a validated model of post-operative growth. This general framework has the potential to be a tool for modeling other complex systems, such as stress-driven growth, fiber-reinforced material formulations, and fluid-structure interactions. It also has the potential to be utilized as a workflow for high-throughput modeling and analysis of population data. Sample source code is provided to enable such development in aortic growth modeling, with the aim to further broaden its applicability and stimulate future translation into clinical work.

## Data Availability

All data produced in the present study are available upon reasonable request to the authors.

## Acknowledgments

We acknowledge the support of the National Institutes of Health (NIH), NHLBI, R01HL159205 (to L.P.). A.D.G. and N.N. acknowledge the partial support of the National Science Foundation (NSF) through grant CMMI-2433223 (to N.N.). The Center for Research Informatics is funded by the Biological Science Division at The University of Chicago with additional funding provided by the Institute for Translational Medicine, CTSA grant number UL1TR002389 from NIH.

## Appendix A

### 3D vs. shell implementations in growth benchmark problems

In this section, we focus on the standard problem of stress-induced hypertension as a benchmark problem for our shell implementation. Previous work has demonstrated the result for 3D solids showing wall thickening to reduce stress back to the homeostatic stress state as a result of the increase in the pressure [17, 40]. Specifically, a neo-Hookean form for *W_e_* is used to determine the growth tensor:

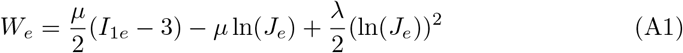

while growth depends on the stress state with the goal of bringing the stress back to the homeostatic state: *F_g_* = *ν̇I*

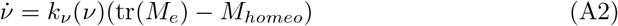

The details of the coefficients *k_ν_* and the input parameters are given in the reference [40]. To further demonstrate the effects of growth laws, we also implemented linear growth.

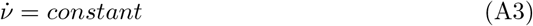

Figure A1 shows the comparisons between the results of three cases for both growth laws. The first is a reference case that uses a standard solver with a 3D formulation (Benchmark) [40], the second is our VUMAT implementation with 3D solid elements (3D: Linear Growth/Stress-Driven Growth), and the third is our VUMAT implementation with shell elements (Shell: Linear Growth/Stress-Driven Growth). As shown in Figure A1d, our 3D VUMAT reproduced the stress and thickness profiles of the reference case. We note a difference between 3D VUMAT and shell VUMAT as a result of the thickness that goes away when the thickness is reduced. These good agreements show that when the thickness of the object is small enough, a shell implementation is reliable and effective.

**Fig. A1.**
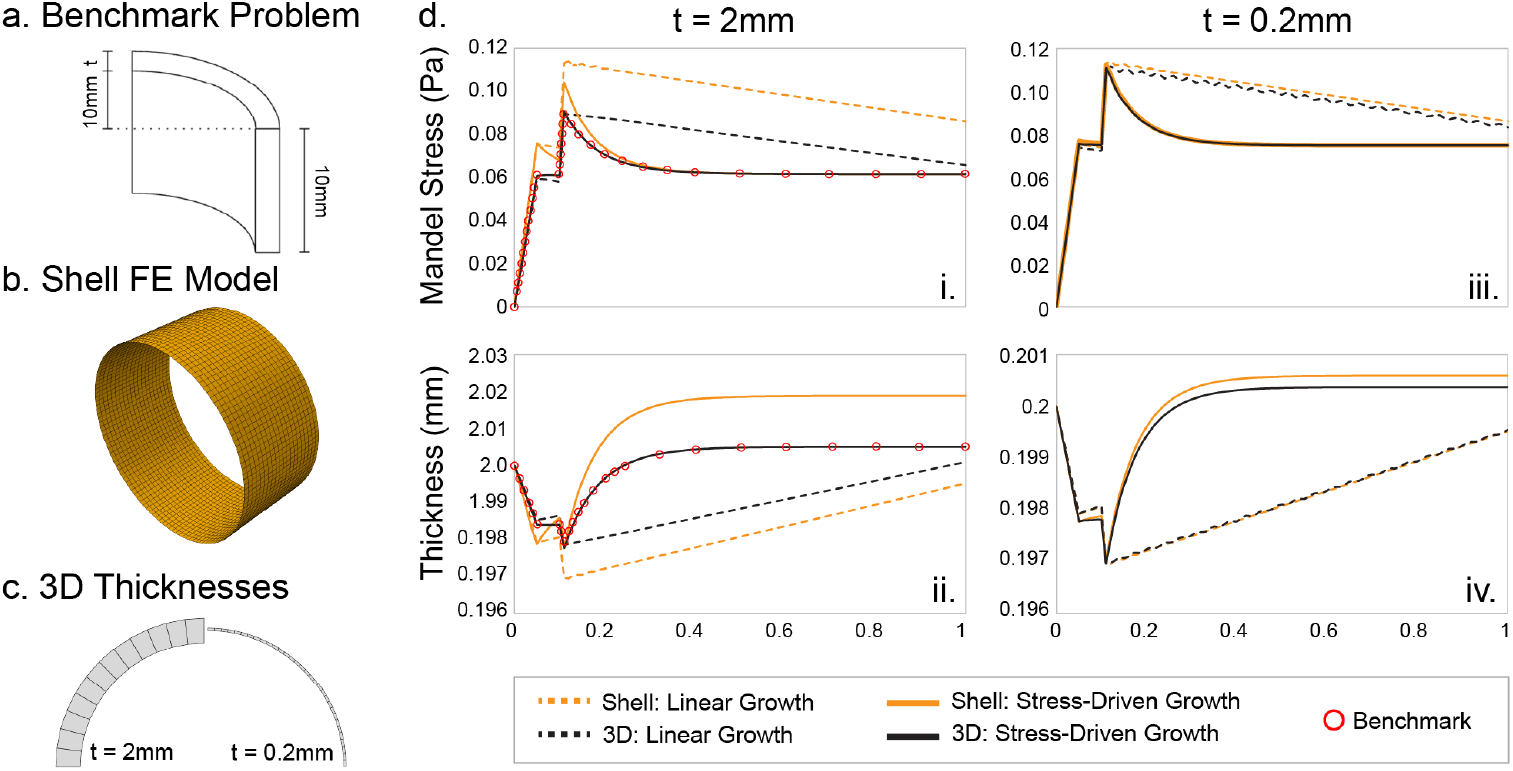
Hypertension benchmark problem, shell replicated 3D growth. a) Dimensions and quarter representation of full geometry of the benchmark problem. b) Representation of the shell finite element (FE) model. c) Depiction of the two thicknesses, 2mm and 0.2mm, for 3D formulation. d) Comparisons of resultant Mandel stress and thickness for thick (2mm, left) and thin (0.2mm, right) cylinders during various growth processes. Linear growth is shown in dashed lines, while stress-driven growth is shown in solid lines. Black lines indicate 3D formulation, yellow lines indicate shell formulation, and red circles show the previously published benchmark result. [39]

## Appendix B

### 3D vs. shell implementations in patient-specific geometry with heterogeneous growth field

The above benchmark problem utilizes very idealized geometry with a homogeneous growth field. Patient-specific geometries are more complex and the growth field can be heterogeneous. In this section, we show both 3D and shell implementations considering these complexities. Previous work has demonstrated that consistent comparisons between them requires structure in through-thickness meshing, utilizing the extruded meshing technique [31] to avoid extremely high number of elements in the 3D model. Here, we will investigate growth in a patient-specific aortic geometry for the 3D imaged-derived growth implementation, followed by a systematic comparison with shell implementation to demonstrate its effectiveness.

**Fig. B2.**
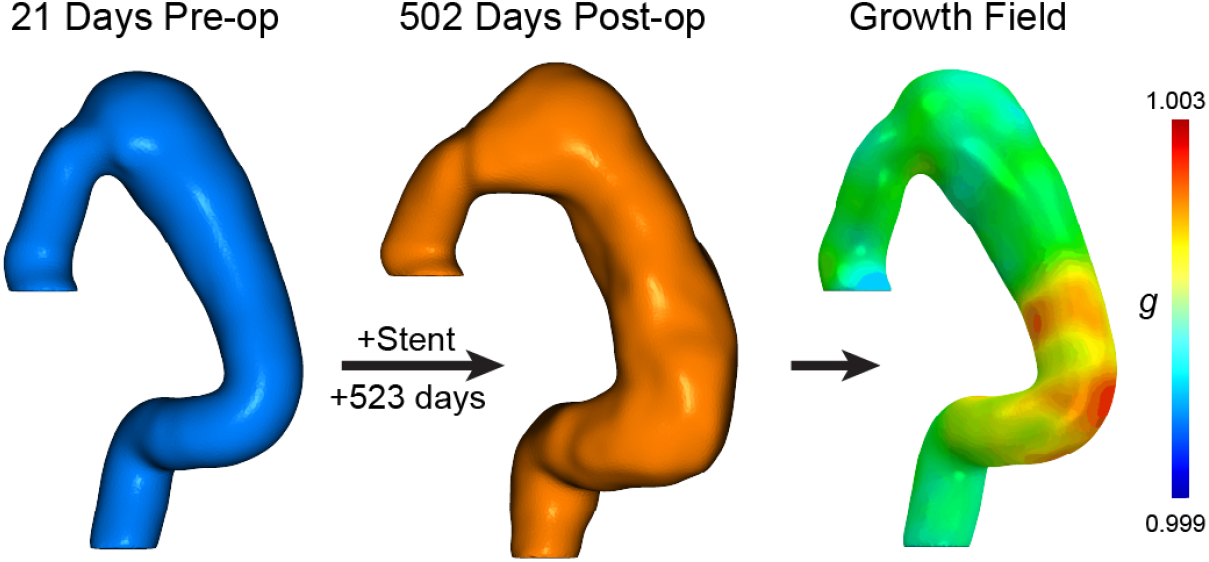
Growth field calculation. Surface segmentations in 3D space of CT image-derived aortic geometry for the patient at 21 days pre-op (blue) and 502 days post-op (orange) are shown. Through registration and calculation of local area changes, the growth field shown on the right is extracted and applied to the models in Figure B3.

Aortic pathology, such as aneurysm and dissection, often leads to significant changes in shape. Figure B2 demonstrates the evolution of a dissected aorta over two timepoints captured by CT imaging. Following the general computational framework established in the main text (Section 2), a heterogeneous growth field is calculated by registration of a patient-specific aortic geometry at 21 days pre-operative and 502 days post-operative, with treatment by stent deployment.

**Fig. B3.**
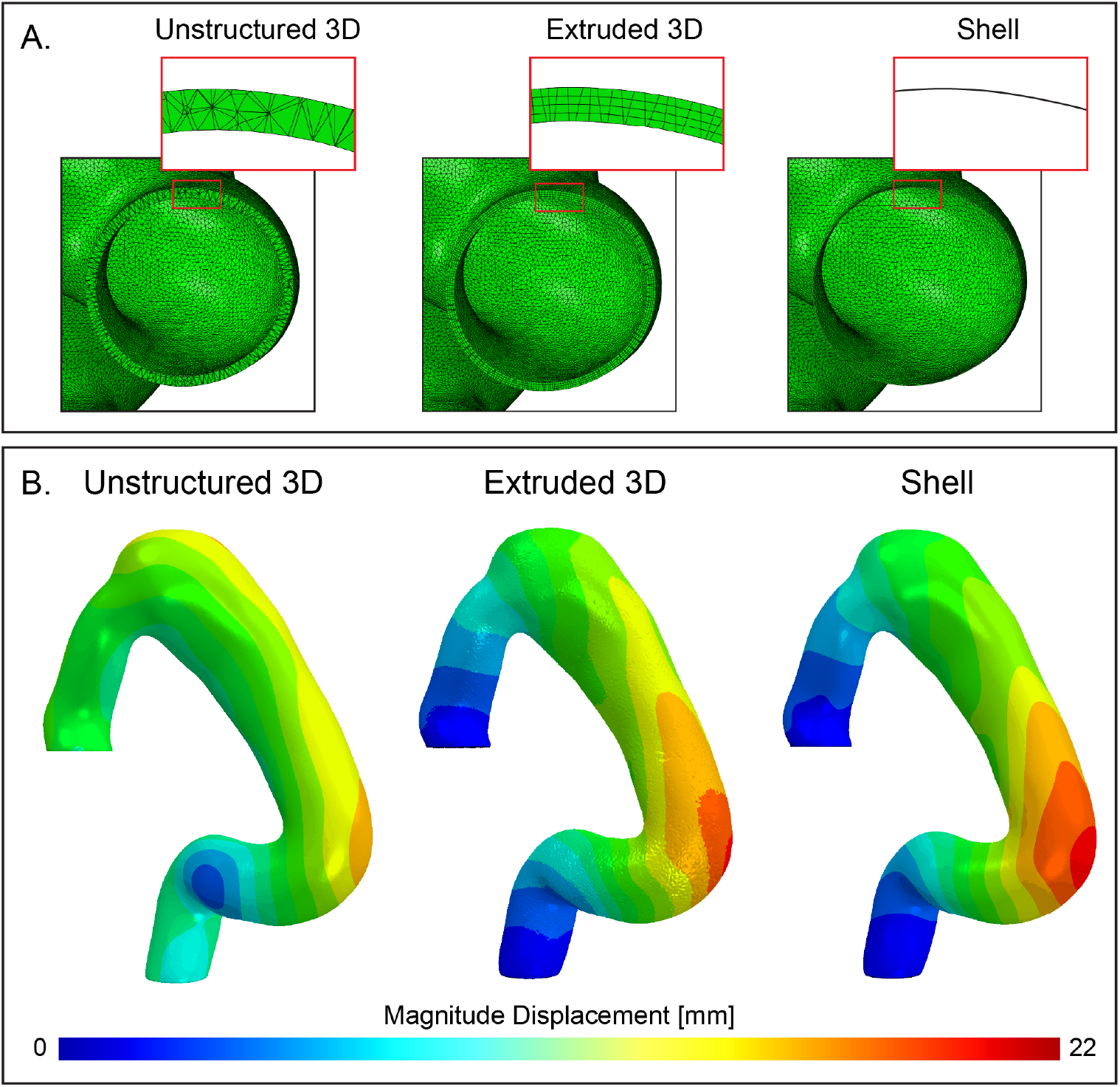
Impact of through-thickness meshing on resultant deformation. (A) Representative cross-sections of each formulation. Notice how the unstructured 3D formulation is composed of both elements that partially and fully span the thickness. The extruded 3D formulation has consistent through-thickness meshing instead. (B) Magnitude displacement fields displayed on the deformed geometries of each model after the implementation of the growth field shown in Figure B2.

The growth field shown in Figure B2 is implemented as described in the main general framework in three formulations: Unstructured 3D, extruded 3D, and shell. The unstructured 3D model is the standard for previous patient-specific growth methodologies [7]. The thickness is segmented and meshed explicitly using a tetrahedral mesh in ScanIp. Errors and noise can arise due to the technical challenge of segmenting the aortic wall and the resolution of the mesh along this thickness. The extruded 3D model instead utilizes a segmented surface and extrudes with an assigned thickness. The extrusion is formed with 4 elements through the thickness. The shell model utilizes a segmented surface with an equivalent thickness assigned to the shell but not explicitly modeled. This shell formulation uses 5 section points, similar to 5 through-thickness nodes of the extruded 3D model. Representative cross-sections of these three models are shown in Figure B3a.

The displacement fields as a result of the growth simulations are exhibited in Figure B3b. Good agreement is shown between the extruded 3D and shell formulations, while the unstructured 3D model shows a very different displacement field. This result indicates that unstructured through-thickness meshing has a strong impact on displacement localization and morphological evolution. While the extruded 3D model shows consistency, highly structured through-thickness meshing is computationally intensive and difficult for patient-specific geometries, especially geometries with more than one branch or cavity, like the dissected aorta modeled in Section 3 of the main text.

## References

1. Sotiras, A., Davatzikos, C., Paragios, N.: Deformable Medical Image Registration: A Survey. IEEE transactions on medical imaging 32(7), 1153 (2013) 10.1109/TMI.2013.2265603 . Accessed 2026-07-19

[2] Burris, N.S., Hoff, B.A., Kazerooni, E.A., Ross, B.D.: Vascular deformation mapping (VDM) of thoracic aortic enlargement in aneurysmal disease and dissection. Tomography 3(3), 163–173 (2017) 10.18383/j.tom.2017.00015

[3] Burris, N.S., Hoff, B.A., Patel, H.J., Kazerooni, E.A., Ross, B.D.: Three-Dimensional Growth Analysis of Thoracic Aortic Aneurysm With Vascular Deformation Mapping. Circulation. Cardiovascular Imaging 11(8), 008045 (2018) 10.1161/CIRCIMAGING.118.008045

[4] Braet, D.J., Eliason, J., Ahmed, Y., Bakel, P.A.J., Zhong, J., Bian, Z., Figueroa, C.A., Burris, N.S.: Vascular Deformation Mapping of Abdominal Aortic Aneurysm. Tomography (Ann Arbor, Mich.) 7(2), 189–201 (2021) 10.3390/tomography7020017

[5] Pugar, J.A., Jiang, D., Kim, J., Pocivavsek, L.: Shape and Scale in Quantifying Aortic Morphology Evolution and Chronicity. Cardiovascular Engineering and Technology 17(3), 343–356 (2026) 10.1007/s13239-026-00827-z . Accessed 2026-07-13

[6] Khabaz, K., Yuan, K., Pugar, J., Jiang, D., Sankary, S., Dhara, S., Kim, J., Kang, J., Nguyen, N., Cao, K., Washburn, N., Bohr, N., Lee, C.J., Kindlmann, G., Milner, R., Pocivavsek, L.: The geometric evolution of aortic dissections: Predicting surgical success using fluctuations in integrated gaussian curvature. PLOS Computational Biology 20(2), 1011815 (2024) 10.1371/journal.pcbi.1011815

[7] Khabaz, K., Kim, J., Milner, R., Nguyen, N., Pocivavsek, L.: Temporal geometric mapping defines morphoelastic growth model of type b aortic dissection evolution. Computers in Biology and Medicine 182, 109194 (2024) 10.1016/j.compbiomed.2024.109194

[8] Taber, L.A.: Biomechanics of Growth, Remodeling, and Morphogenesis. Applied Mechanics Reviews 48(8), 487–545 (1995) 10.1115/1.3005109 . Accessed 2026-07-17

[9] Rodriguez, E.K., Hoger, A., McCulloch, A.D.: Stress-dependent finite growth in soft elastic tissues. Journal of Biomechanics 27(4), 455–467 (1994) 10.1016/0021-9290(94)90021-3

[10] Skalak, R., Zargaryan, S., Jain, R.K., Netti, P.A., Hoger, A.: Compatibility and the genesis of residual stress by volumetric growth. Journal of Mathematical Biology 34(8), 889–914 (1996) 10.1007/BF01834825

[11] Ambrosi, D., Ateshian, G.A., Arruda, E.M., Cowin, S.C., Dumais, J., Goriely, A., Holzapfel, G.A., Humphrey, J.D., Kemkemer, R., Kuhl, E., Olberding, J.E., Taber, L.A., Garikipati, K.: Perspectives on biological growth and remodeling. Journal of the Mechanics and Physics of Solids 59(4), 863–883 (2011) 10.1016/j.jmps.2010.12.011

12. Himpel, G., Kuhl, E., Menzel, A., Steinmann, P.: Computational Modelling of Isotropic Multiplicative Growth. Computer Modeling in Engineering & Sciences 8(2), 119–134 (2005) 10.3970/cmes.2005.008.119 . Publisher: Tech Science Press. Accessed 2026-07-18

[13] Humphrey, J.D.: Remodeling of a collagenous tissue at fixed lengths. Journal of Biomechanical Engineering 121(6), 591–597 (1999) 10.1115/1.2800858 https://asmedigitalcollection.asme.org/biomechanical/article-pdf/121/6/591/5766986/5911.pdf

[14] Humphrey, J.D., Rajagopal, K.R.: A constrained mixture model for arterial adaptations to a sustained step change in blood flow. Biomechanics and Modeling in Mechanobiology 2(2), 109–126 (2003) 10.1007/s10237-003-0033-4

15. Valentín, A., Humphrey, J.D., Holzapfel, G.A.: A finite element-based constrained mixture implementation for arterial growth, remodeling, and adaptation: theory and numerical verification. International Journal for Numerical Methods in Biomedical Engineering **29**(8), 822–849 (2013) 10.1002/cnm.2555

[16] Cyron, C.J., Aydin, R.C., Humphrey, J.D.: A homogenized constrained mixture (and mechanical analog) model for growth and remodeling of soft tissue. Biomechanics and modeling in mechanobiology 15(6), 1389 (2016) 10.1007/s10237-016-0770-9 . Accessed 2026-07-22

17. Kuhl, E., Maas, R., Himpel, G., Menzel, A.: Computational modeling of arterial wall growth: Attempts towards patient-specific simulations based on computer tomography. Biomechanics and Modeling in Mechanobiology 6(5), 321–331 (2007) 10.1007/s10237-006-0062-x

[18] Watton, P.N., Hill, N.A., Heil, M.: A mathematical model for the growth of the abdominal aortic aneurysm. Biomechanics and Modeling in Mechanobiology 3(2), 98–113 (2004) 10.1007/s10237-004-0052-9

[19] Watton, P.N., Hill, N.A.: Evolving mechanical properties of a model of abdominal aortic aneurysm. Biomechanics and Modeling in Mechanobiology 8(1), 25–42 (2009) 10.1007/s10237-007-0115-9

20. Fereidoonnezhad, B., Naghdabadi, R., Holzapfel, G.A.: Stress softening and permanent deformation in human aortas: Continuum and computational modeling with application to arterial clamping. Journal of the Mechanical Behavior of Biomedical Materials **61**, 600–616 (2016) 10.1016/j.jmbbm.2016.03.026

21. Fereidoonnezhad, B., Naghdabadi, R., Sohrabpour, S., Holzapfel, G.A.: A Mechanobiological model for damage-induced growth in arterial tissue with application to in-stent restenosis. Journal of the Mechanics and Physics of Solids 101, 311–327 (2017) 10.1016/j.jmps.2017.01.016 . Publisher: Pergamon. Accessed 2026-07-18

[22] Sáez, P., Peña, E., Martìnez, M.A., Kuhl, E.: Computational modeling of hypertensive growth in the human carotid artery. Computational Mechanics 53(6), 1183–1196 (2014) 10.1007/s00466-013-0959-z

23. Cardamone, L., Valentín, A., Eberth, J.F., Humphrey, J.D.: Modelling carotid artery adaptations to dynamic alterations in pressure and flow over the cardiac cycle. Mathematical Medicine and Biology: A Journal of the IMA 27(4), 343–371 (2010) 10.1093/imammb/dqq001

[24] Lashkarinia, S.S., Coban, G., Kose, B., Salihoglu, E., Pekkan, K.: Computational modeling of vascular growth in patient-specific pulmonary arterial patch reconstructions. Journal of Biomechanics 117, 110274 (2021) 10.1016/j.jbiomech.2021.110274

[25] Taylor, C.A., Figueroa, C.A.: Patient-specific modeling of cardiovascular mechanics. Annual Review of Biomedical Engineering 11(1), 109–134 (2009) 10.1146/annurev.bioeng.10.061807.160521

[26] Sala, A., Vincentiis, C.d., Grimaldi, F., Rubino, B., Cirami, M., Perillo, N., Vitale, R., Cardani, R., Boveri, S., Conti, M., Spagnolo, P.: Can the novel photon-counting CT scan accurately predict aortic wall thickness? preliminary results. Bioengineering 12(3) (2025) 10.3390/bioengineering12030306

[27] Chiastra, C., Migliori, S., Burzotta, F., Dubini, G., Migliavacca, F.: Patient-specific modeling of stented coronary arteries reconstructed from optical coherence tomography: Towards a widespread clinical use of fluid dynamics analyses. Journal of Cardiovascular Translational Research 11(2), 156–172 (2018) 10.1007/s12265-017-9777-6

[28] Curcio, N., Rosato, A., Mazzaccaro, D., Nano, G., Conti, M., Matrone, G.: 3d patient-specific modeling and structural finite element analysis of atherosclerotic carotid artery based on computed tomography angiography. Scientific Reports 13(1), 19911 (2023) 10.1038/s41598-023-46949-5

[29] Gnyaneshwar, R., Kumar, R.K., Balakrishnan, K.R.: Dynamic analysis of the aortic valve using a finite element model. The Annals of Thoracic Surgery 73(4), 1122–1129 (2002) 10.1016/s0003-4975(01)03588-3

[30] Nama, N., Aguirre, M., Ortigosa, R., Gil, A.J., Humphrey, J.D., Figueroa, C.A.: A systematic comparison between membrane, shell, and 3d solid formulations for non-linear vascular biomechanics. arXiv (2023) 10.48550/arxiv.2301.10504 2301.10504

[31] Kim, T., Ortigosa, R., Nama, N., Aguirre, M., Gil, A.J., Humphrey, J.D., Figueroa, C.A.: A systematic comparison of membrane, shell, and 3d solid formulations for nonlinear vascular biomechanics. Journal of the Mechanical Behavior of Biomedical Materials 179, 107423 (2026) 10.1016/j.jmbbm.2026.107423

[32] Noble, C., Carlson, K.D., Neumann, E., Dragomir-Daescu, D., Erdemir, A., Lerman, A., Young, M.: Patient specific characterization of artery and plaque material properties in peripheral artery disease. Journal of the Mechanical Behavior of Biomedical Materials 101, 103453 (2020) 10.1016/j.jmbbm.2019.103453

[33] Straughan, R., Kadry, K., Parikh, S.A., Edelman, E.R., Nezami, F.R.: Fully automated construction of three-dimensional finite element simulations from optical coherence tomography. ArXiv (2024)

[34] Laubrie, J.D., Mousavi, J.S., Avril, S.: A new finite-element shell model for arterial growth and remodeling after stent implantation. International Journal for Numerical Methods in Biomedical Engineering 36(1), 3282 (2020) 10.1002/cnm.3282

[35] Rolf-Pissarczyk, M., Schussnig, R., Fries, T.-P., Fleischmann, D., Elefteriades, J.A., Humphrey, J.D., Holzapfel, G.A.: Mechanisms of aortic dissection: From pathological changes to experimental and in silico models. Progress in Materials Science 150, 101363 (2025) 10.1016/j.pmatsci.2024.101363

[36] Vorp, D.A., Raghavan, M.L., Muluk, S.C., Makaroun, M.S., Steed, D.L., Shapiro, R., Webster, M.W.: Wall strength and stiffness of aneurysmal and nonaneurysmal abdominal aorta. Annals of the New York Academy of Sciences 800(1), 274–276 (1996) 10.1111/j.1749-6632.1996.tb33330.x

[37] Raghavan, M.L., Vorp, D.A.: Toward a biomechanical tool to evaluate rupture potential of abdominal aortic aneurysm: identification of a finite strain constitutive model and evaluation of its applicability. Journal of Biomechanics 33(4), 475–482 (2000) 10.1016/S0021-9290(99)00201-8

[38] Donik, Dhara, S., Li, W., Nnate, B., Sankary, S., Polcari, K., Varsanik, M.A., Khabaz, K., Milner, R., Nguyen, N., Kramberger, J., Pocivavsek, L.: Bare metal stenting for residual arch dissections: A computational analysis. Cardiovascular Engineering and Technology 16(6), 624–640 (2025) 10.1007/s13239-025-00799-6

39. Tinkl, S.: Towards a predictive computational growth model for aneurysms. PhD thesis, Technische Universität München (2015). https://mediatum.ub.tum.de/1230842

[40] Sáez, P., Peña, E., Martínez, M.A., Kuhl, E.: Computational modeling of hypertensive growth in the human carotid artery. Computational mechanics 53(6), 1183 (2013) 10.1007/s00466-013-0959-z . Accessed 2026-07-18

